# Evaluating Diabetes Drug Targets for Vascular Dementia Prevention Using Drug Target Mendelian Randomisation

**DOI:** 10.64898/2026.09.22.26363326

**Authors:** Amy Packer, Lauren Austin, Dylan M. Williams, Victoria Taylor-Bateman, Emma L. Anderson

## Abstract

Vascular dementia (VaD) is a leading cause of cognitive decline with no disease-modifying therapies. It remains unclear whether diabetes drug targets causally influence VaD risk. We applied two-sample, drug target Mendelian randomisation (MR) to test whether modulation of eight antidiabetic drug targets may alter VaD risk. MR models used variants within genes encoding antidiabetic drug targets, selected according to their effects on circulating glycated haemoglobin (HbA1c, N = 437,749) in primary analyses, and cortex-specific *cis*-eQTLs (N = 2,683) in secondary analyses. Outcomes were clinically diagnosed VaD (N = 7,009 cases/899,672 controls) and neuroimaging markers of cerebrovascular disease: white matter hyperintensity volume (WMH; N = 50,970), inverse fractional anisotropy (iFA; N = 31,125), and mean diffusivity (MD; N = 31,147). Type-2 diabetes mellitus (T2DM; N= 62,892 cases/596,424 controls) served as a positive control outcome. Genetic instruments were available for seven of the relevant targets using both HbA1c and cortex *cis*-eQTL data. In primary HbA1c-scaled analyses, no antidiabetic drug target showed evidence of association with VaD risk or cerebrovascular neuroimaging markers, although *DPP4* inhibition demonstrated a suggestive protective association for VaD (OR = 0.68, 95% CI = 0.46 - 1.02, p = 0.063), though the confidence interval (CI) included the null. Secondary cortex *cis*-eQTL analyses were similarly null, except for *PPARG* modulation, which was predicted to reduce iFA (β = −0.50, 95% CI −0.99 to −0.02, p = 0.04). Overall, these findings provide little genetic evidence supporting currently licensed antidiabetic drugs as candidates for vascular dementia prevention, although *DPP4* inhibition warrants further investigation in larger datasets.

## Introduction

More than an estimated 57 million people are living with dementia worldwide (Nichols et al., 2022). Vascular dementia (VaD) is the second most common cause (Bir et al., 2021), yet no pharmacological therapies are currently approved specifically for its treatment (Lennon & Sachdev, 2026). Drug repurposing offers an efficient strategy to accelerate therapeutic development (Cummings et al., 2025), but applications to VaD remain limited (Taylor-Bateman et al., 2026).

Type 2 diabetes mellitus (T2DM) is a proposed modifiable risk factor for dementia (Livingston et al., 2024) and has been associated with increased risk of VaD in observational studies (Wang et al., 2023). Mendelian randomisation (MR) studies suggest that T2DM may contribute to VaD risk and some cerebral small vessel disease phenotypes (Litkowski et al., 2023; J. Liu et al., 2018). However, findings across cerebrovascular outcomes are inconsistent, and analyses of genetically predicted HbA1c have reported null associations (J. Liu et al., 2018; Taylor-Bateman et al., 2022). Relatedly, several antidiabetic drug classes, including sodium-glucose cotransporter 2 (SGLT2) inhibitors, thiazolidinediones (TZDs), glucagon-like peptide-1 (GLP-1) receptor agonists, and dipeptidyl peptidase-4 (DPP-4) inhibitors, have been associated with lower dementia risk (Li et al., 2024; Tang et al., 2023). However, evidence remains inconsistent (Kuate Defo et al., 2024; Stefanou et al., 2026). Observational studies remain susceptible to residual confounding (Boyko, 2013; Eberly et al., 2021; Kornblith et al., 2022), while randomised controlled trial (RCT) evidence is limited and inconclusive (Li et al., 2024; Lin et al., 2023). Further, most trials are small and restricted to older or metabolically impaired populations, limiting assessment of effects on prodromal cerebrovascular pathology (d’Arbeloff et al., 2019). Consequently, the effects of diabetes therapies on VaD remain uncertain.

Drug target MR leverages genetic variants near genes encoding therapeutic targets to study the effects of pharmacological target modulation on disease outcomes (Gill et al., 2021). This approach can help prioritise candidates for drug repurposing, as therapeutic targets supported by human genetic evidence are approximately twice as likely to gain regulatory approval as those without such evidence (King et al., 2019). Limited research has employed drug target MR to evaluate antidiabetic drugs for VaD. To our knowledge, only one drug target MR study has evaluated genetically proxied GLP-1 receptor agonism in relation to VaD and markers of cerebral small vessel disease, reporting lower risk of small vessel stroke and lower white matter hyperintensity volume (Zangas et al., 2026). However, its less stringent instrument selection strategy may have increased susceptibility to genetic confounding. Consequently, the repurposing potential of diabetes drug targets for VaD remains uncertain.

We therefore used a two-sample drug target MR design to investigate whether genetically proxied modulation of eight diabetes drug targets influences risk of clinically diagnosed VaD and neuroimaging markers of cerebrovascular pathology.

## Materials and Methods

### Study overview

A two-sample, drug target MR framework (also known as *cis*-MR) was employed to evaluate whether modulation of diabetes drug targets influences the risk of VaD (Sanderson et al., 2022). For the primary analysis, *cis*-acting single nucleotide polymorphisms (SNPs) for each T2DM drug target were identified from a genome-wide association study (GWAS) of glycated haemoglobin (HbA1c) (Barton et al., 2021), a downstream biomarker indicative of target modulation across T2DM drugs. Genetic association data for these SNPs were then extracted from our outcome GWAS datasets for the study outcomes. T2DM was included as a positive control outcome for instrument validation (Xue et al., 2018). Four VaD-related outcomes comprised clinical VaD diagnosis (Taylor-Bateman et al., 2026) and neuroimaging markers of cerebrovascular pathology: white matter hyperintensity volume (WMH) (Sargurupremraj et al., 2020), fractional anisotropy (FA) (Taylor-Bateman et al., 2022), and mean diffusivity (MD) (Taylor-Bateman et al., 2022). Secondary analyses used cortex-specific expression quantitative trait loci (eQTLs) (de Klein et al., 2023) for each target, instead of HbA1c. A flowchart outlining the main stages of the study is presented in Figure 1.

**Figure 1.**
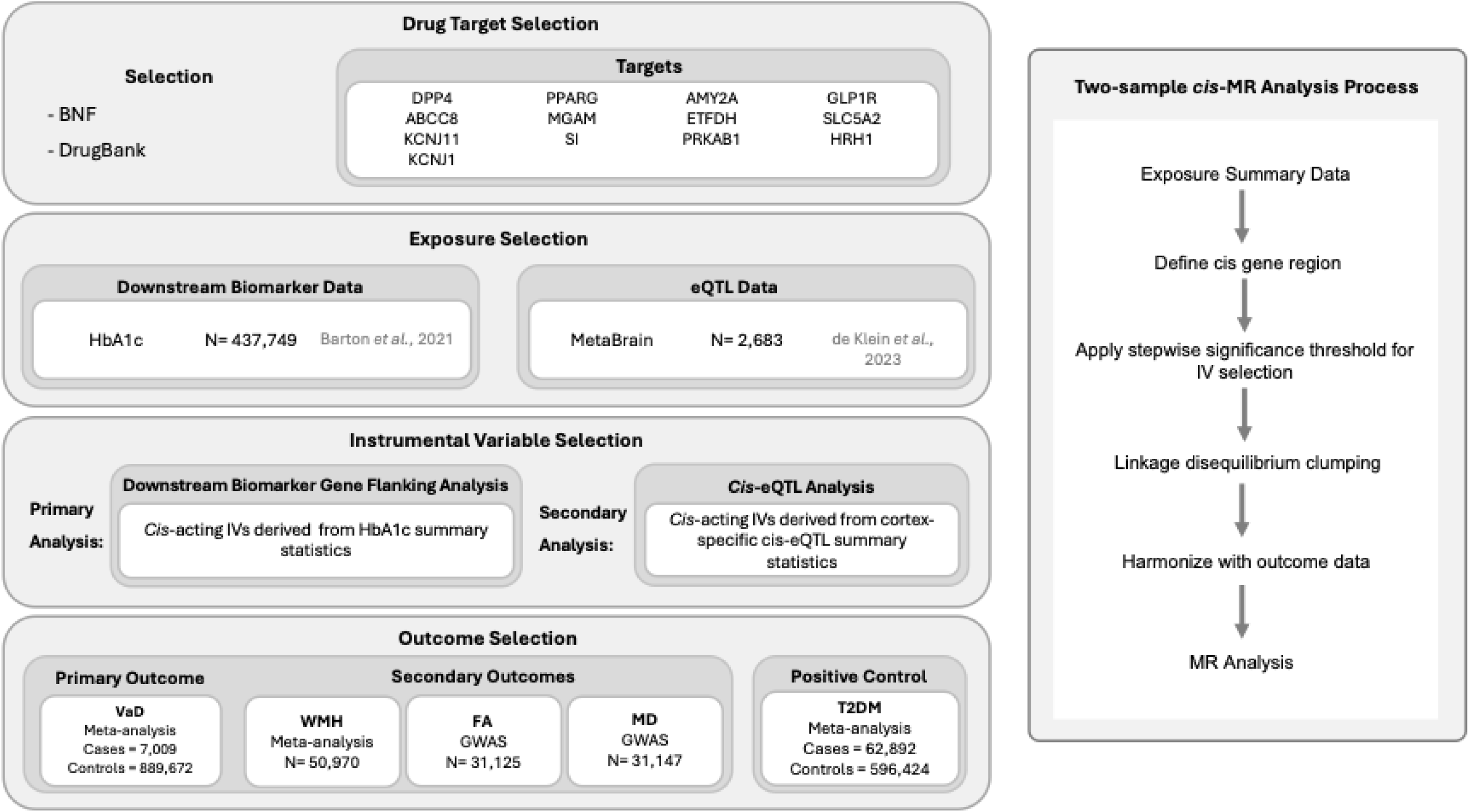
Study Design and Analytic Framework for Drug Target Mendelian Randomisation of Antidiabetic Agents and Vascular Dementia. Antidiabetic agents were identified through the BNF (*BNF Content Published by NICE*, 2026) and their corresponding molecular targets obtained from DrugBank (Knox et al., 2024). IVs were selected from GWAS summary statistics for HbA1c (Barton et al., 2021) to proxy target modulation using a downstream biomarker (primary analysis), and from cortex-specific *cis*-eQTL data (de Klein et al., 2023) to proxy target gene expression (secondary analysis). *Cis*-acting single nucleotide polymorphisms (SNPs) were included at genome-wide significance or p < 1 × 10⁻⁵ if no genome-wide significant variants were available. SNPs were pruned for linkage disequilibrium prior to harmonisation with outcome data. The primary outcome was clinically defined VaD (Taylor-Bateman et al., 2026). Secondary outcomes were MRI markers of cerebrovascular pathology: WMH (Sargurupremraj et al., 2020), FA (Taylor-Bateman et al., 2022), and MD (Taylor-Bateman et al., 2022). T2DM was included as a positive control outcome to validate instrument selection. Abbreviations: BNF = British National Formulary; HbA1c = glycated haemoglobin; eQTL = expression quantitative trait loci; GWAS = genome-wide association study; IV = instrument variable; VaD = vascular dementia; WMH = white matter hyperintensities; FA = fractional anisotropy; MD = mean diffusivity; T2DM = type 2 diabetes mellitus; MR = Mendelian randomisation.

### Selection of Diabetes Drug Targets

Agents licensed for the treatment of T2DM in the United Kingdom (UK) were identified through the British National Formulary (BNF) (*BNF Content Published by NICE*, 2026). Restricting analyses to UK-licensed therapies was intended to maximise the translational relevance of the findings. Drug classes included biguanides, GLP-1 receptor agonists, DPP-4 inhibitors, SGLT2 inhibitors, TZDs, sulfonylureas, α-glucosidase inhibitors, and meglitinides. Pharmacological targets, corresponding to antidiabetic agents with defined mechanisms of action, were obtained from DrugBank (Knox et al., 2024), and genomic regions for the genes encoding these targets were defined using the National Centre for Biotechnology Information (NCBI) Gene database (Sayers et al., 2025). Gene coordinates and the *cis*-regions used for instrument selection are provided in Supplementary Table 1.

Eight unique drug targets were identified and included in analyses (Table 1). For drug classes with a single shared target, this representative target was used in analyses: GLP-1 receptor agonists (*GLP1R*), DPP-4 inhibitors (*DPP4*), and SGLT2 inhibitors (*SLC5A2*). For drug classes with multiple recognised targets, target-specific effects of drugs in those classes were evaluated separately: sulfonylureas (*ABCC8*, *KCNJ11*, *KCNJ1*), TZDs (*PPARG*), and meglitinides (*ABCC8*, *HRH1*).

**Table 1.**
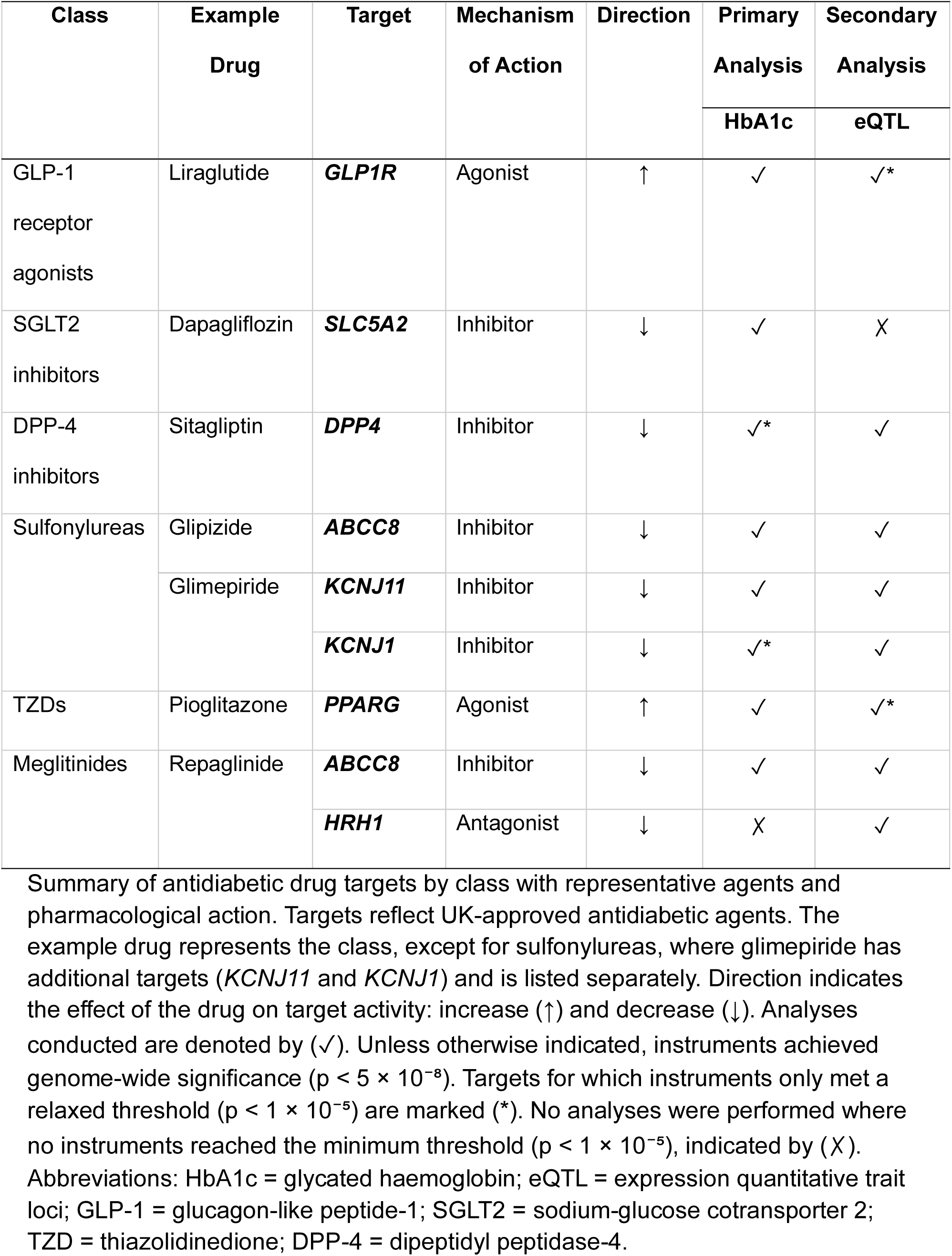
Summary of Identified Targets and Corresponding Analyses Performed.

Metformin (biguanide) and acarbose (α-glucosidase inhibitor) were excluded because they lack clearly defined pharmacological targets that can be reliably genetically proxied. Metformin exerts its effects through multiple incompletely characterised pathways and putative targets, several of which remain under debate (Anderson & Williams, 2023; Rena et al., 2017), whereas acarbose acts on a family of α-glucosidase enzymes rather than a clearly defined target protein, limiting the feasibility of a target-based Mendelian randomisation approach.

### MR Assumptions

MR estimates are valid under the instrumental variable (IV) assumptions of relevance (IV1), independence from confounders (IV2), and absence of horizontal pleiotropy (IV3; Figure 2). Supplementary Box 1 outlines these assumptions and summarises the steps taken to minimise and evaluate potential violations.

**Figure 2.**
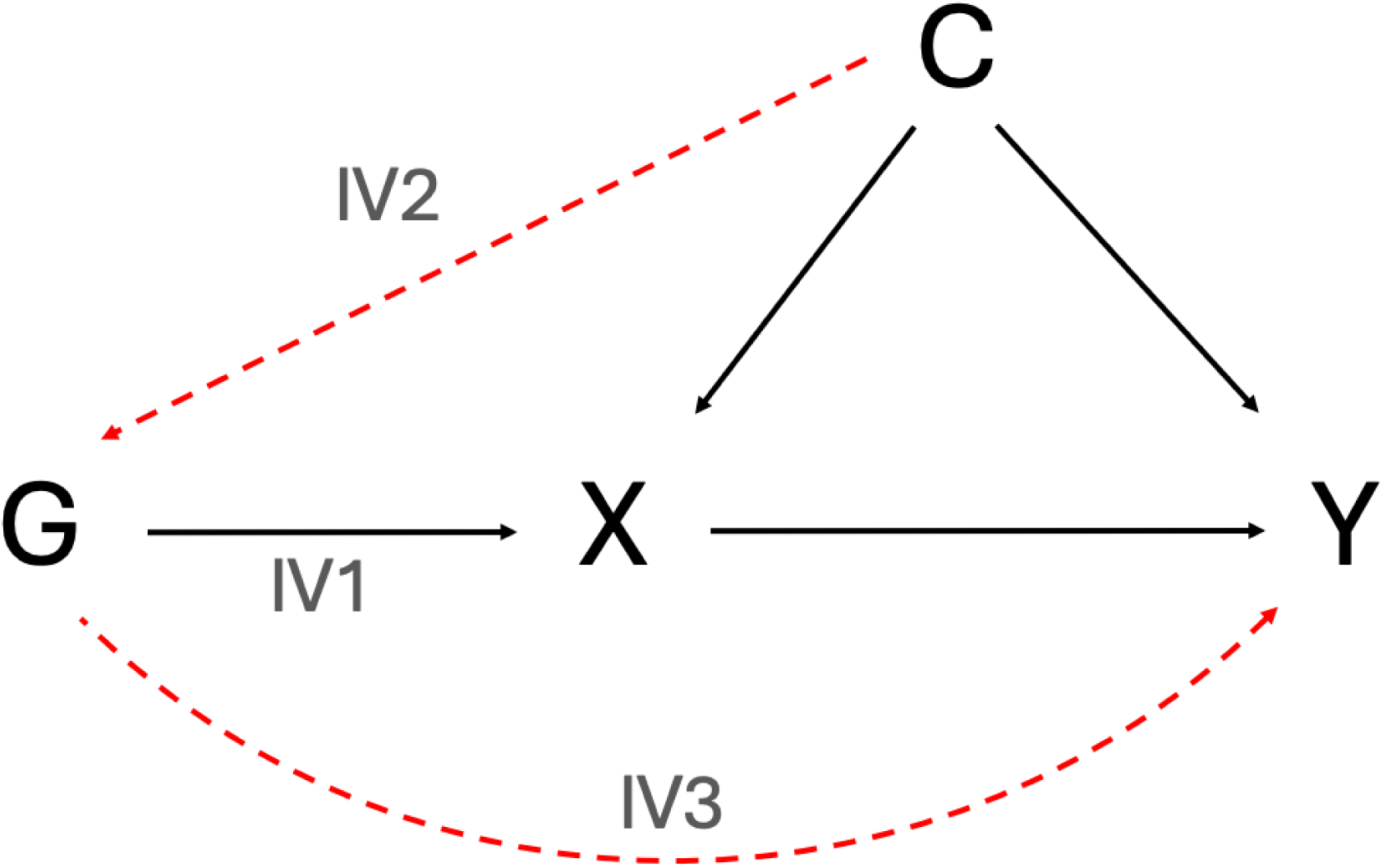
Core Assumptions of Mendelian Randomisation. G denotes a genetic variant, X the exposure, Y the outcome, and C confounders. IV1: Genetic variants used as IVs must be robustly associated with the exposure. IV2: No confounding of the variant-outcome relationship. IV3: Variants influence the outcome only through the exposure and not alternative biological pathways.

### Data

All GWAS summary statistics were derived from individuals of European ancestry to minimise population stratification and confounding (i.e. violation of IV2). For each exposure and outcome, the largest available GWAS dataset was used to maximise statistical power. All summary-level data were publicly available. Ethical approval and participant consent for each contributing cohort were obtained in the original studies, as reported in the corresponding publications. An overview of all summary-level data is provided in Table 2.

**Table 2.** Datasets Used for Exposure and Outcome in Mendelian Randomisation Analyses.

| **GWAS** | **Purpose** | **Source** | **Sample Size** | **Cohorts** | **Data Ascertainment Method(s)** |
| --- | --- | --- | --- | --- | --- |
| **HbA1c** | Exposure | Barton et al., 2021 (Barton et al., 2021) | 437,749 | UK Biobank (Bycroft et al., 2018) | Whole blood samples via HPLC |
| **Cortex-specific eQTL** | Exposure | de Klein et al., 2023 (de Klein et al., 2023) | 2,683 | AMP-AD consortium (Hodes & Buckholtz, 2016); Braineac (Ramasamy et al., 2014); PsychENCODE consortium (Akbarian et al., 2015); BrainSeq (Schubert et al., 2015); NABEC (Gibbs et al., 2010); TargetALS (Prudencio et al., 2015); GTEx (Lonsdale et al., 2013); ENA (Leinonen et al., 2011). | Post-mortem RNA-sequencing |
| **VaD** | Outcome | Taylor-Bateman et al., 2026<br>(Taylor-Bateman et al., 2026) | 7,009/<br>899,672 | MEGACVID (The Mega Vascular Cognitive Impairment and Dementia (MEGAVCID) consortium, 2024); FinnGen (Kurki et al., 2023) | Electronic health records, ICD-10 codes, clinical assessment |
| **WMH** | Outcome | Sargurupremraj et al., 2020<br>(Sargurupremraj et al., 2020) | 50,970 | CHARGE (Psaty et al., 2009); UK Biobank (Bycroft et al., 2018) | Brain MRI (T2-weighted FLAIR), automated segmentation of WMH volume |
| **FA** | Outcome | Taylor-Bateman et al., 2022<br>(Taylor-Bateman et al., 2022) | 31,125 | UK Biobank (Bycroft et al., 2018) | Brain MRI using DTI sequences with automated processing |
| **MD** | Outcome | Taylor-Bateman et al., 2022<br>(Taylor-Bateman et al., 2022) | 31,147 | UK Biobank (Bycroft et al., 2018) | Brain MRI using DTI sequences with automated processing |
| **T2DM** | Positive<br>Control | Xue et al., 2018 (Xue et al., 2018) | 62,892/<br>596,424 | DIAGRAM (Morris et al., 2012), GERA (Banda et al., 2015), UK Biobank (Bycroft et al., 2018) | Clinical diagnosis |

The table summarises the genome-wide association study (GWAS) datasets used as sources of summary-level data for exposures and outcomes for Mendelian randomisation (MR) analyses. The UK Biobank contributed to both exposure and outcome datasets. Abbreviations: HPLC = high-performance liquid chromatography; AMP-AD = Accelerating Medicines Partnership – Alzheimer’s Disease; NABEC = North American Brain Expression Consortium; GTEx = Genotype-Tissue Expression Project; ENA = European Nucleotide Archive; FLAIR = fluid-attenuated inversion sequences; DIAGRAM; Diabetes Genetics Replication and Meta-analysis Consortium; GERA = Genetic Epidemiology Research on Adult Health and Aging cohort; MEGACVID = Mega Vascular Cognitive Impairment and Dementia; CHARGE = Cohorts for Heart and Aging Research in Genetic Epidemiology; DTI = diffusion tensor imaging; HbA1c = glycated haemoglobin; eQTL = expression quantitative trait loci; VaD = vascular dementia; WMH = white matter hyperintensities; FA = fractional anisotropy; MD = mean diffusivity; T2DM = type 2 diabetes mellitus; MRI = magnetic resonance imaging; RNA = ribonucleic acid; ICD-10 = International Classification of Diseases, 10th revision.

#### Exposure data

##### Primary analysis: Targets Scaled to Effect on HbA1c

HbA1c measured in whole blood served as a downstream biomarker of drug target function for *cis*-acting variants in the gene regions encoding each target, scaling genetic instruments as per expected pharmacological modulation of the targets. HbA1c provides an integrated measure of mean glycaemia over the preceding 8-12 weeks and is routinely used for the diagnosis and monitoring of T2DM (Mehta et al., 2025). Summary-level data for HbA1c were obtained from a GWAS of 437,749 UK Biobank participants (Barton et al., 2021), a large prospective population-based cohort of adults aged 37–73 years at baseline assessments in 2006–2010 (Sudlow et al., 2015). Prior to association testing, HbA1c values underwent quality control, covariate adjustment, and inverse normal transformation. Consequently, SNP-HbA1c association estimates are expressed in standard deviation units of transformed HbA1c.

##### Secondary analysis: Cortex-specific cis-eQTLs

As a secondary analysis, cortex-specific *cis*-eQTLs were incorporated to instrument genetically predicted expression of target genes in cortical tissue, allowing interrogation of tissue-relevant effects and providing a complementary approach to the primary HbA1c-scaled analyses. Summary-level genome-wide *cis*-eQTL data were obtained from the cortex-specific MetaBrain consortium (N = 2,683) (de Klein et al., 2023).

#### Outcome Data

For the positive control outcome analysis, summary-level GWAS data for T2DM risk were drawn from a meta-analysis of 62,892 cases and 596,424 (Xue et al., 2018). Given that the selected agents are approved for the treatment of T2DM, genetic modulation of their corresponding targets should also be associated with T2DM risk, provided sufficient statistical power. Evidence of a significant risk reduction supports the relevance and validity of the instruments. Where variants were unavailable in the T2DM dataset, proxy variants (r^2^ > 0.6) were sought using the LDlink proxy tool (Machiela & Chanock, 2015), selecting the first available variant with the highest r^2^ value. Full details of proxy selection methods and SNPs are available in the Supplementary Materials. Supplementary Table 2 displays the original SNPs and proxies used.

The primary outcome was clinically diagnosed VaD. Summary-level data were obtained from a VaD case-control GWAS meta-analysis, yielding a total sample of 7,009 cases and 899,672 controls (Taylor-Bateman et al., 2026). The meta-analysis comprised data from the Mega Vascular Cognitive Impairment and Dementia (MEGAVCID) consortium and the FinnGen study (Kurki et al., 2023; The Mega Vascular Cognitive Impairment and Dementia (MEGAVCID) consortium, 2024). Structural MRI markers, WMH (N=50,970) (Sargurupremraj et al., 2020), FA (N = 31,125) (Taylor-Bateman et al., 2022), and MD (N = 31,147) (Taylor-Bateman et al., 2022), were evaluated as additional outcomes. To ensure consistent interpretability across outcomes, FA was directionally harmonised by inverting its values (iFA), such that higher values across all traits uniformly reflect poorer cerebrovascular measures. The three MRI markers were selected due to their well-established associations with vascular cognitive impairment and their ability to sensitively detect subclinical cerebrovascular pathology (Debette et al., 2019; Pasi et al., 2016). Unlike binary clinical endpoints such as VaD risk, these continuous measures provide greater statistical power to detect smaller effects of drug target modulation.

### Statistical Analyses

All analyses were performed using R (version 4.4.3). For each diabetes drug target, a *cis*-region extending ±500 kb around the gene coding sequence was defined to minimise the risk of horizontal pleiotropy (Supplementary Table 1). SNPs within this region were retained if they met genome-wide significance (p < 5 × 10⁻⁸) with the exposure trait (HbA1c or eQTLs); if no genome-wide significant SNPs were available, a relaxed threshold of p < 1 × 10⁻⁵ was applied. To ensure independence among instruments, linkage disequilibrium (LD) clumping was performed using a 10,000 kb window and r² < 0.001 based on the 1000 Genomes EUR reference panel (The 1000 Genomes Project Consortium et al., 2015). The clumping p-value threshold (p₁) was matched to the SNP selection threshold applied for each analysis (p < 5 × 10⁻⁸ for genome-wide significance, or p < 1 × 10⁻⁵ where no genome-wide significant variants were available). For each drug target, a separate set of instruments was generated using this procedure from summary-level HbA1c and cortex-specific cis-eQTL data.

IVs were harmonised with outcome data to align effect alleles between exposure and outcome datasets. Allele frequency information was used to infer strand orientation for palindromic variants where possible, while ambiguous palindromic variants for which strand orientation could not be determined reliably were excluded. MR estimates were based on the Wald ratio (Sanderson et al., 2022) method for targets instrumented by a single SNP, and the inverse-variance weighted (IVW) method for targets with multiple instruments (Sanderson et al., 2022). Analyses were conducted using the *TwoSampleMR* (Hemani et al., 2018) and *MendelianRandomization* (Yavorska & Burgess, 2017) R packages. As all targets contained fewer than 3 instruments after data curation steps, sensitivity analyses that require 3 or more IVs were precluded. Instrument strength (IV1) was evaluated using F-statistics, with values over 10 taken to indicate sufficient relevance and low risk of weak-instrument bias (Burgess et al., 2011). Heterogeneity across variants (IV3) was evaluated using Cochran’s Q statistic when multiple instruments were available (Bowden et al., 2015).

Results are reported as odds ratios (ORs) for binary outcomes and mean unit differences for continuous outcomes. Primary analyses were scaled per 0.1-SD genetically predicted reduction in transformed HbA1c attributable to target modulation, with estimates oriented to reflect the glucose-lowering action of diabetes drugs. Secondary analyses were scaled per 1-SD genetically predicted change in target gene expression, with effect directions aligned to the expected therapeutic action of each diabetes drug target (agonism, antagonism, or inhibition), ensuring genetically-proxied changes reflected the drugs’ mechanisms. In figures, log-odds were plotted instead of ORs for binary outcomes to facilitate scaling when plotting results for binary outcomes alongside continuous ones.

No formal multiple-testing correction was applied. Bonferroni correction would be overly stringent in this setting, given multiple related drug targets across multiple related outcomes, raising potential false negatives. Instead, associations were interpreted cautiously, with careful consideration of confidence intervals and triangulation of supporting evidence.

### Reporting guidelines

This study followed the Strengthening the Reporting of Observational Studies in Epidemiology Using Mendelian Randomisation (STROBE-MR) guidelines (Skrivankova, Richmond, Woolf, Davies, et al., 2021; Skrivankova, Richmond, Woolf, Yarmolinsky, et al., 2021), with the checklist included in the Supplementary Materials.

## Results

Genetic instruments were identified for seven of the eight selected diabetes drug targets in the primary HbA1c-scaled analyses; no suitable instrument was available for *HRH1*. In the secondary cortex-specific cis-eQTL analyses, instruments were available for seven targets, with no suitable instrument available for *SLC5A2* (Table 1). The genetic variants used as instrumental variables are recorded in Supplementary Table 3 (HbA1c analyses) and Supplementary Table 4 (cortex-specific *cis*-eQTL analyses). Additionally, some MR analyses could not be conducted due to loss of instruments during harmonisation or absence of SNPs in the outcome GWAS. In *cis*-eQTL analyses, this applied to *HRH1* for VaD, iFA, and MD, and to *GLP1R* for VaD and WMH.

In HbA1c-scaled analyses, proxy variants were used for positive control analyses for *DPP4* (rs6733162 (r² = 0.876)) and *KCNJ1* (rs12797048 (r² = 0.611)) (Supplementary Table 2). In the *cis*-eQTL analyses, a proxy variant was used for positive control analyses for *KCNJ1* (rs948215, (r² = 0.673); however, positive control testing could not be performed for *DPP4* and *GLP1R*, as no suitable proxy variants were identified in the outcome dataset.

Across both primary and secondary analyses, negative effect estimates indicate associations in the direction of benefit, corresponding to lower T2DM or VaD risk, or to better brain health for neuroimaging outcomes. Effect estimates from HbA1c-scaled and cortex-specific *cis*-eQTL-analyses are not directly comparable, as they reflect different scales of target perturbation (i.e., genetically predicted HbA1c reduction versus genetically predicted target gene expression). Full MR results are presented in full in the Supplementary Table 5 (HbA1c analyses) and Supplementary Table 6 (cortex-specific *cis*-eQTL analyses).

### Primary Analysis: Targets Scaled to Effect on HbA1c

In the positive control outcome analyses, reduced T2DM risk was predicted for modulation of *GLP1R* (OR = 0.85, 95% CI = 0.77 - 0.94, p = 0.001), *ABCC8* (OR = 0.79, 95% CI = 0.75 - 0.84, p < 0.001), *KCNJ11* (OR = 0.79, 95% CI = 0.75 - 0.84, p < 0.001), and *DPP4* (OR = 0.78, 95% CI = 0.66 - 0.92, p = 0.003), confirming that these are likely valid for downstream MR analyses (Figure 3; Supplementary Table 5). In contrast, there was no clear evidence for differences in T2DM risk from MR models that indexed modulation of *PPARG*, *KCNJ1* and *SLC5A2* (Supplementary Figure 1; Supplementary Table 5). Thus, downstream results for VaD and neuroimaging biomarkers for *PPARG*, *KCNJ1* and *SLC5A2* should be interpreted more cautiously than results for targets validated in the positive control outcome analyses and are therefore presented separately. All instruments demonstrated sufficient strength (F-statistics > 10). The only analysis using multiple SNP instruments (*SLC5A2* and WMH in the HbA1c-scaled analyses) showed no evidence of heterogeneity (Cochran’s Q, p > 0.050), indicating consistent SNP-specific estimates.

**Figure 3.**
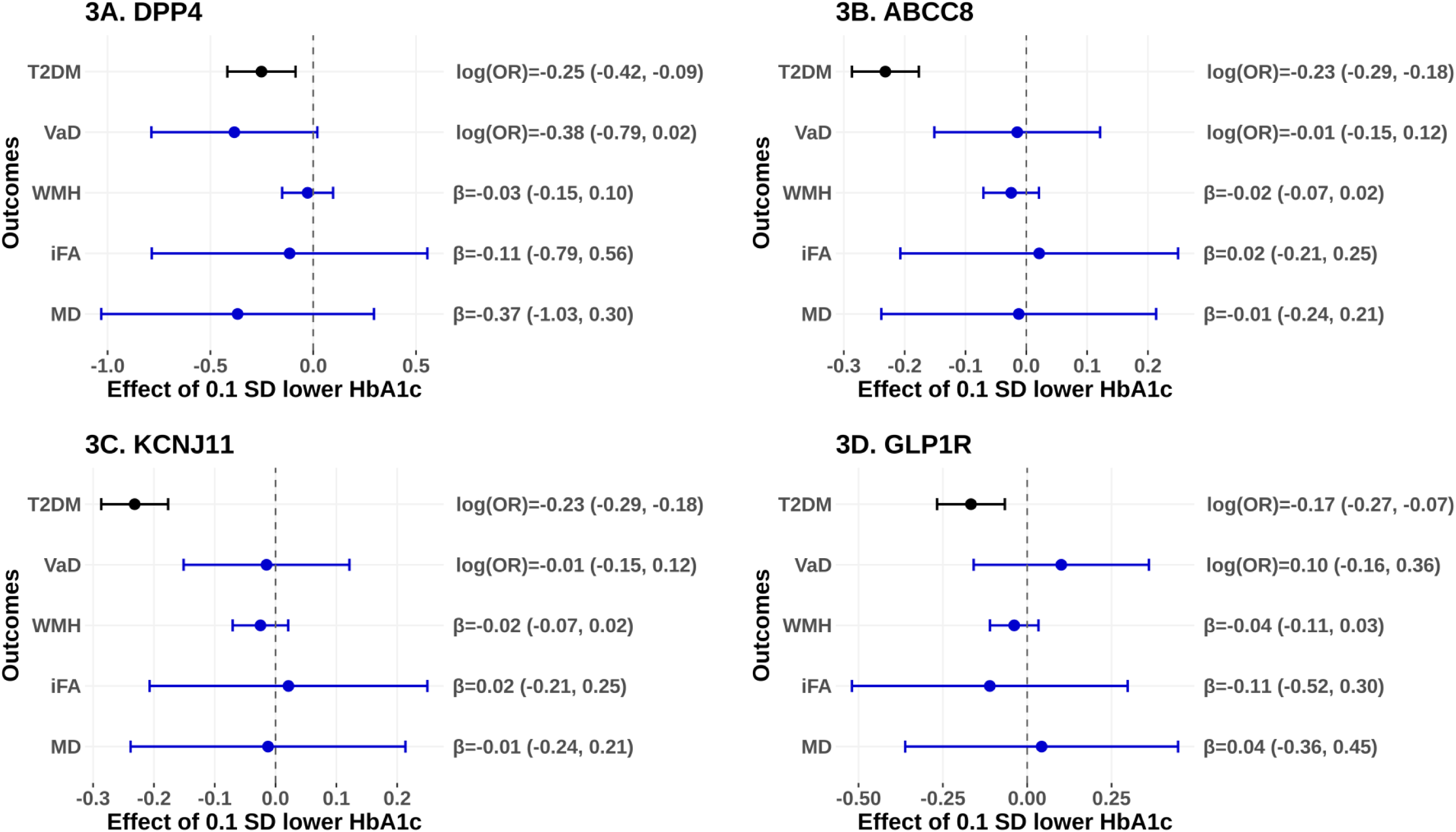
Primary MR estimates of effects of modulating antidiabetic drug targets (*DPP4*, *ABCC8*, *KCNJ11*, *GLP1R*) on T2DM risk and vascular dementia-related outcomes. Results for the following targets are displayed as follows: (A) *DPP4*, (B) *ABCC8*, (C) *KCNJ11*, and (D) *GLP1R* – all of which were validated by the positive control outcome analyses depicted for T2DM. Effect estimates represent the change associated with a 0.1-SD decrease in genetically instrumented HbA1c. β coefficients are plotted for continuous outcomes (WMH, iFA, MD), whereas log-odds are reported for binary outcomes (VaD and T2DM). Point estimates and corresponding 95% confidence intervals are displayed to the right of each plot. F-statistics exceeded 10 in all analyses, indicating adequate instrument strength. The *ABCC8* (B) and *KCNJ11* (C) analyses both used the same single SNP as the genetic instrument. Abbreviations: T2DM = type 2 diabetes mellitus; VaD = vascular dementia; WMH = white matter hyperintensities; iFA = inverse fractional anisotropy; MD = mean diffusivity; SD = standard deviation; HbA1c = glycated haemoglobin; MR = Mendelian randomisation.

In the HbA1c-scaled analysis of target effects on cerebrovascular outcomes, no clear evidence of an association with VaD risk or neuroimaging outcomes (WMH, iFA, MD) was identified (Figure 3; Supplementary Figure 1; Supplementary Table 5). A suggestive association was observed for *DPP4* in relation to VaD risk, where genetically predicted inhibition was linked to reduced risk of VaD (OR = 0.68, 95% CI = 0.46 - 1.02, p = 0.063). However, this estimate was imprecise with wide confidence intervals spanning the null.

### Secondary Analysis: Cis-eQTLs

Full results from the secondary analyses, using cortex-specific *cis*-eQTLs, are reported in the Supplementary Results and Supplementary Table 6. Overall, there was also limited evidence that modulation of these drug targets would be expected to influence VaD risk or neuroimaging outcomes. *PPARG* modulation was estimated to reduce iFA (β = −0.50, 95% CI −0.99 to −0.02, p = 0.04) but estimates for other cerebrovascular outcomes all centred on the null (Supplementary Figure 1). There were contradictory predicted directions of effect of *GLP1R* agonism on iFA (β= 0.36, 95% CI 0.05 to 0.72, p = 0.05) and MD (β= -0.33, 95% CI -0.68 to 0.03, p = 0.07) (Supplementary Figure 2); though these results should be interpreted cautiously since it was not possible to test the indexing of *GLP1R* agonism with eQTL data against T2DM risk for positive control outcome validation.

## Discussion

In this two-sample, drug target MR analysis, we found little evidence that modulation of currently licensed antidiabetic drug targets influences VaD risk or related neuroimaging outcomes - although a suggestive finding for *DPP4* inhibition on VaD (combined with a lack of precision for that result) would warrant follow-up in future research.

These findings add to the inconclusive evidence regarding the potential role of glucose-lowering therapies in dementia prevention (Kuate Defo et al., 2024; Li et al., 2024; Lin et al., 2023; Stefanou et al., 2026). Our estimates reflect genetically proxied target modulation in a general population sample and therefore provide evidence that is complementary to, but distinct from, observational and clinical studies. To our knowledge, only one previous drug target MR study has evaluated an antidiabetic drug target in relation to VaD and related cerebrovascular phenotypes. That study reported that genetically proxied GLP-1 receptor agonism was associated with lower risk of small vessel stroke and reduced WMH volume (Zangas et al., 2026). However, a less stringent instrument selection strategy was used than in the present study, which may have increased susceptibility to genetic confounding. In contrast, our more conservative instrument selection yielded little evidence that modulation of *GLP1R*, or other currently licensed antidiabetic drug targets, substantially influences VaD risk or related cerebrovascular phenotypes, including WMH volume.

Although our findings provided little overall support for antidiabetic drug targets in VaD prevention, *DPP4* inhibition emerged as a potential exception, showing a consistent direction of association across outcomes and suggestive evidence of lower VaD risk. DPP-4 is a widely expressed proteolytic enzyme found in plasma and cerebrospinal fluid, with biological functions extending beyond glucose regulation, including roles in inflammatory, immune, and vascular signalling pathways (Klemann et al., 2016; Mulvihill & Drucker, 2014; Ribeiro-Silva et al., 2023). While these findings warrant further investigation, the imprecision of the estimates with confidence intervals encompassing the null precludes firm conclusions. Replication using larger, better-powered VaD genetic datasets will be required to establish whether *DPP4* inhibition truly influences disease risk.

Overall, our findings provide little genetic evidence supporting currently licensed antidiabetic drug targets as priorities for vascular dementia prevention. These results are informative for therapeutic prioritisation because they suggest that several widely discussed diabetes drug targets are unlikely to exert large protective effects on VaD risk, while identifying *DPP4* inhibition as a target warranting further investigation.

### Strengths and limitations

Key strengths of this study include the application of two-sample drug target Mendelian randomisation to reduce bias from confounding and reverse causation, and the use of large-scale genetic datasets to maximise statistical power. The complementary HbA1c-scaled and cortex-specific *cis*-eQTL analyses expanded the range of available targets and enabled triangulation of evidence. Further, positive control analyses and neuroimaging endophenotypes provided additional validation and triangulation of evidence. Importantly, genetically informed null findings are valuable for therapeutic prioritisation because they can identify targets that are unlikely to yield clinically meaningful benefits, thereby reducing investment in costly and lengthy dementia prevention trials with a low probability of success.

Several limitations warrant consideration. First, a key limitation is that our results reflect target-specific effects of perturbing specific molecular targets, rather than the effects of administering the corresponding drug. For agents acting predominantly through a single target (e.g., *GLP1R*), these estimates may reasonably approximate pharmacological effects. However, for drugs with multiple targets or where there are relevant off-target effects of therapeutic agents, the estimated effects may not encompass all aspects of exposure to drugs of interest. Consequently, findings should be interpreted as target-specific effects rather than estimates of drug use. Further, genetic variants proxy lifelong perturbation of drug targets and therefore most closely resemble the effects of long-term target modulation. These estimates may not correspond to the effects of pharmacological treatment initiated later in life or administered over shorter durations. This distinction may be particularly relevant for targets where the biological consequences of chronic perturbation differ from those of acute pharmacological modulation. Second, there were some instrument-related limitations. For some targets, relaxed p-value thresholds were required to identify instruments, potentially compromising IV1 and increasing horizontal pleiotropy risk (IV3). Some instruments were additionally absent from the T2DM GWAS, limiting positive-control validation. Targets were instrumented with one or two SNPs, precluding pleiotropy-robust sensitivity analyses such as MR-Egger. However, by using only *cis*-acting variants, the risk of pleiotropy is greatly minimised. Additionally, although cortex-specific eQTLs enhance tissue relevance, gene–trait associations may still partly reflect expression effects in unrelated tissues (de Klein et al., 2023) (a potential violation of IV3). Moreover, RNA expression does not necessarily reflect protein levels or activity (Y. Liu et al., 2016). Brain protein QTLs (pQTLs) would provide more direct proxies of the most potentially relevant pharmacological perturbation, but brain pQTL resources remain limited in scale relative to available eQTL datasets. Third, as SNP-exposure estimates for HbA1c, FA, and MD were obtained from the discovery GWAS, effect sizes may therefore be inflated due to Winner’s curse, biasing MR estimates towards the null (Jiang et al., 2023). Overall, these caveats largely reflect structural data limitations rather than weaknesses in study design. Fourth, restricting analyses to European ancestry likely reduced bias from population stratification (minimising potential violations of IV2; Supplementary Box 1). However, the generalisability of findings to other ancestries remains uncertain. Future analyses should incorporate ancestry-specific and cross-ancestry comparisons as larger multi-ancestry GWAS become available. Finally, there was partial sample overlap between exposure and outcome GWAS for some analyses, which may bias estimates and inflate Type I error rates (Burgess et al., 2016). However, given adequate instrument F-statistics throughout, any bias is likely to be modest (Burgess et al., 2016; Sadreev et al., 2021).

### Future directions

Collectively, these findings help refine therapeutic target prioritisation for VaD prevention. By providing human genetic evidence against major effects of several currently licensed antidiabetic drug targets, this study may help focus future repurposing efforts on more promising therapeutic mechanisms. *DPP4* inhibition remains a plausible candidate and should be evaluated in larger genetic studies and complementary research designs.

## Data Availability

All summary statistics were obtained from publicly available datasets or directly from the corresponding authors, with details from the original papers as described in Table 2, refs. (Barton et al., 2021; de Klein et al., 2023; Sargurupremraj et al., 2020; Taylor-Bateman et al., 2022, 2026; Xue et al., 2018).

