## Supplementary Materials for "Evaluating Diabetes Drug Targets for Vascular Dementia Prevention Using Drug Target Mendelian Randomisation"

##### Methods

##### MR Assumptions

| Mendelian Randomisation Assumption | How assumption was addressed | Tests |
| --- | --- | --- |
| **IV1:** genetic variants used as instruments must be robustly associated with the exposure | Instrumental variables were selected at genome-wide significance threshold where possible | F-statistic was calculated to test for weak instruments ($F < 10$) |
| **IV2:** there must be no confounding between genetic variants and the outcome. | Summary data was obtained from the same ancestral group minimising potential confounding by population stratification |  |
| **IV3:** variants must influence the outcome solely through the exposure and not via alternative biological pathways | Only *cis*-acting instruments were used as they have clearly plausible biological mechanisms, minimising the risk of pleiotropic effects | Heterogeneity across variants was evaluated using Cochran's Q statistic |

*Supplementary Box 1: How assumptions were addressed and evaluation of Mendelian randomisation instrumental assumptions in this study.*

##### Datasets

The VaD diagnosis GWAS was a meta-analysis combining data from MEGACVID (The Mega Vascular Cognitive Impairment and Dementia (MEGAVCID) consortium, 2024), a large international consortium comprising 21 cohorts and consortia, including the UK Biobank (Bycroft et al., 2018). VaD phenotypes were harmonised across cohorts using validated diagnostic criteria, International Classification of Diseases, 10th Revision (ICD-10) codes, clinical assessments, and electronic health records. The FinnGen study (Kurki et al., 2023) is a nationwide Finnish biobank initiative that integrates nationwide health registry data with genome-wide genotyping.

The cortex specific eQTL data was obtained from MetaBrain (de Klein et al., 2023). This initiative integrates post-mortem ribonucleic (RNA) sequencing and genotyping data across 14 cohorts, alongside publicly available datasets archived in the European Nucleotide Archive, including the Accelerating Medicines Partnership – Alzheimer’s Disease (AMP-AD) consortium (Hodes & Buckholtz, 2016), Braineac (Ramasamy et al., 2014), PsychENCODE (Akbarian et al., 2015), BrainSeq (Schubert et al., 2015), North American Brain Expression Consortium (NABEC) (Gibbs et al., 2010), TargetALS (Prudencio et al., 2015), Genotype-Tissue Expression Project (GTEx) (Lonsdale et al., 2013), and European Nucleotide Archive (ENA) (Leinonen et al., 2011).

Summary-level genome-wide association data for WMH volume were obtained from a meta-analysis of 50,970 individuals (Sargurupremraj et al., 2020), including 24,182 participants from the Cohorts for Heart and Aging Research in Genetic Epidemiology (CHARGE) consortium (Psaty et al., 2009) and 26,788 from the UK Biobank (Bycroft et al., 2018). Summary-level GWAS statistics for FA (N = 31,125) (Taylor-Bateman et al., 2022) and MD (N = 31,147) (Taylor-Bateman et al., 2022) were obtained from imaging-derived phenotypes within the UK Biobank (Bycroft et al., 2018). T2DM summary data (Xue et al., 2018) was obtained from the Diabetes Genetics Replication and Meta-analysis (DIAGRAM) consortium (Morris et al., 2012), Genetic Epidemiology Research on Adult Health and Aging (GERA) cohort (Banda et al., 2015), and UK Biobank, with T2DM based on clinical records.

#### **Proxy SNP Analysis**

Where an exposure SNP was unavailable in the T2DM outcome GWAS, proxy variants were identified using the LDlink *LDproxy* tool based on the European (EUR) 1000 Genomes reference panel to match the ancestry of the outcome dataset. For each missing variant, linkage disequilibrium (LD) proxies were searched within the same genomic region and ranked according to pairwise LD ($r^2$) with the original SNP. We prespecified selection of the proxy with the highest available $r^2$. Proxies with $r^2 \geq 0.6$ were accepted due to restriction to *cis*-acting instruments. Proxies were required for *DPP4*, *SLC5A2*, and *KCNJ1* in the HbA1c analyses, and for *DPP4*, *GLP1R*, and *KCNJ1* in the cortex-specific *cis*-eQTL analyses. Supplementary Table 2 displays the original SNPs and proxies used where suitable proxies were identified.

### **Results**

#### **Secondary Analysis: *Cis*-eQTLs**

In the secondary analysis using cortex-specific *cis*-eQTLs, effect estimates were oriented to reflect the pharmacological action of each agent. Specifically, decreases in gene expression were modelled for targets of inhibitory or antagonistic agents, whereas increases in gene expression were modelled for targets of agonistic agents. (See Table 1).

In the positive control analysis for T2DM, increased cortical *PPARG* expression was associated with reduced diabetes risk (OR = 0.76, 95% CI = 0.68 - 0.85, $p < 0.001$, 1 SNP), supporting the validity of this instrument. Similarly, decreased cortical expression of *KCNJ11* was linked to lower T2DM risk (OR = 0.80, 95% CI = 0.75 - 0.85, $p < 0.001$; 1 SNP), further confirming instrument validity. In contrast, decreased expression of *ABCC8* was associated with an increased risk of T2DM (OR = 1.07, 95% CI = 1.03 - 1.10, $p < 0.001$, 1 SNP), opposite to the expected direction of effect. All other instruments demonstrated sufficient strength (F-stat > 10). All analyses used only one SNP as an instrument, precluding assessment of heterogeneity. Supplementary Table 4 shows the genetic variants used as instrumental variables in the secondary analyses, and full results for all targets and outcomes are presented in Supplementary Table 6.

Overall, there was limited evidence that modulation of cortical gene expression at these loci influenced VaD risk or neuroimaging outcomes (WMH, iFA, MD). Outcome-specific associations were identified for *PPARG* and *GLP1R*. A 1-SD increase in cortical *PPARG* expression was associated with reduced iFA ($\beta = -0.50$, 95% CI = -0.99 - -0.02, $p = 0.041$) (Supplementary Figure 2a). In contrast, a 1-SD increase in cortical expression of *GLP1R* was associated with increased iFA ($\beta = 0.36$, 95% CI = 0.01 - 0.73, $p = 0.047$) (Supplementary Figure 2c). Only *PPARG* showed an association with the positive control

#### 1A. PPARG

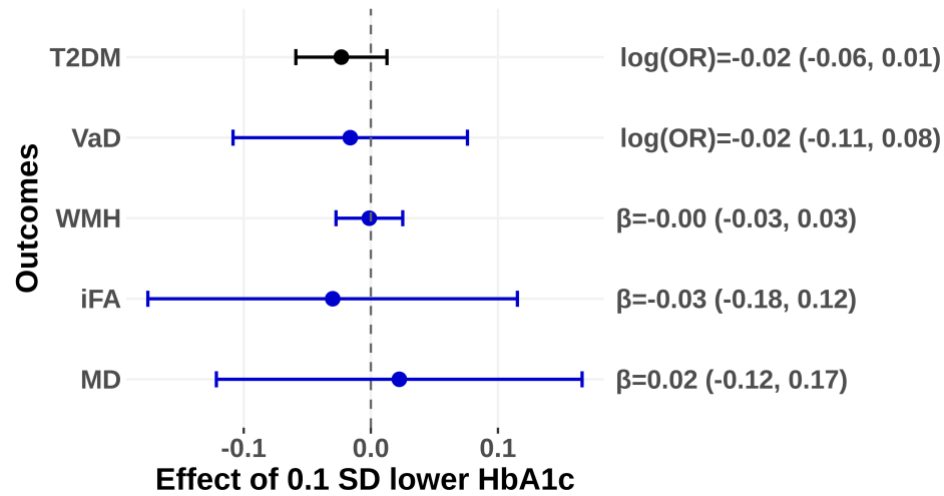

#### 1B. KCNJ1

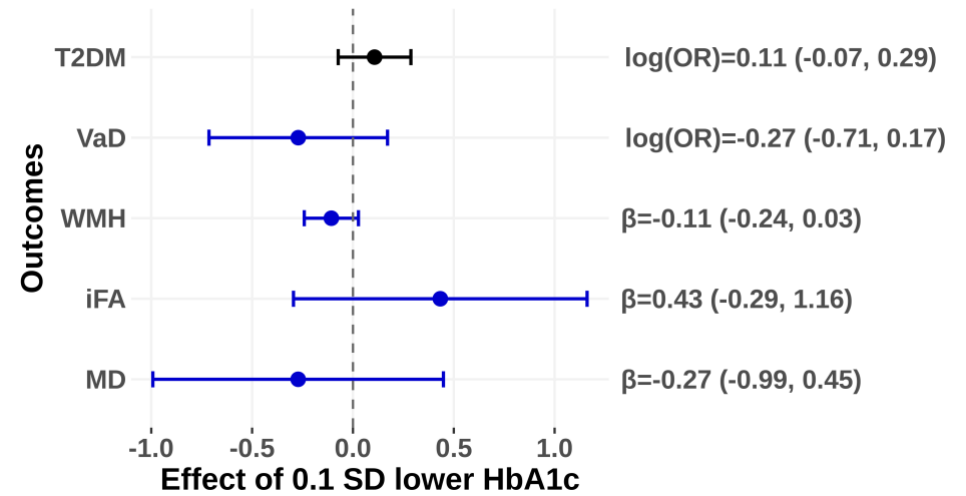

#### 1C. SLC5A2

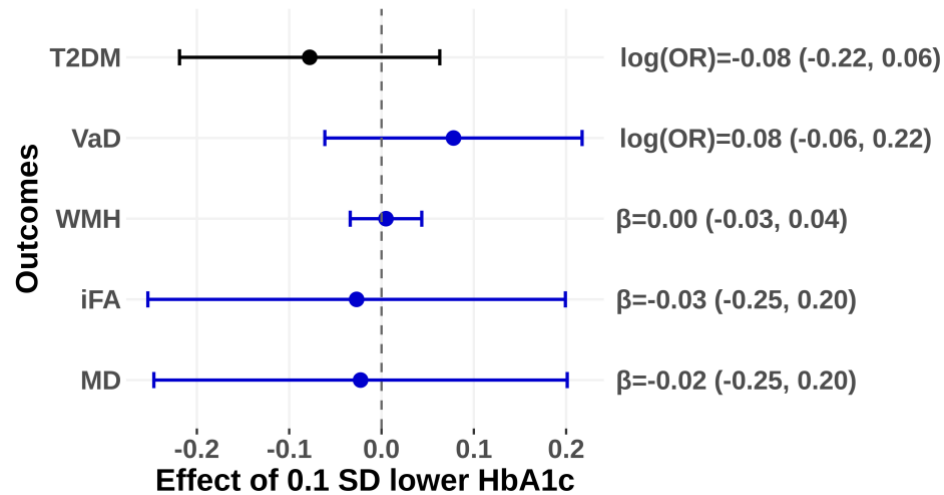

**Supplementary Figure 1. Primary MR estimates of effects of modulating antidiabetic drug targets (*PPARG*, *KCNJ1*, *SCL5A2*) on T2DM risk and vascular dementia-related outcomes**

Results for the following targets are displayed as follows: (A) *PPARG*, (B) *KCNJ1*, and (C) *SCL5A2* – all of which had non-confirmatory results from positive control analyses for T2DM, as depicted. Effect estimates represent the change associated with a 0.1-SD decrease in genetically instrumented HbA1c. $\beta$ coefficients are plotted for continuous outcomes (WMH, iFA, MD), whereas log-odds are reported for binary outcomes (VaD and T2DM). Point estimates and corresponding 95% confidence intervals are displayed to the right of each plot. F-statistics exceeded 10 in all analyses, indicating adequate instrument strength. Abbreviations: T2DM = type 2 diabetes mellitus; VaD = vascular dementia; WMH = white matter hyperintensities; iFA = inverse fractional anisotropy; MD = mean diffusivity; SD = standard deviation; MR = Mendelian randomization.

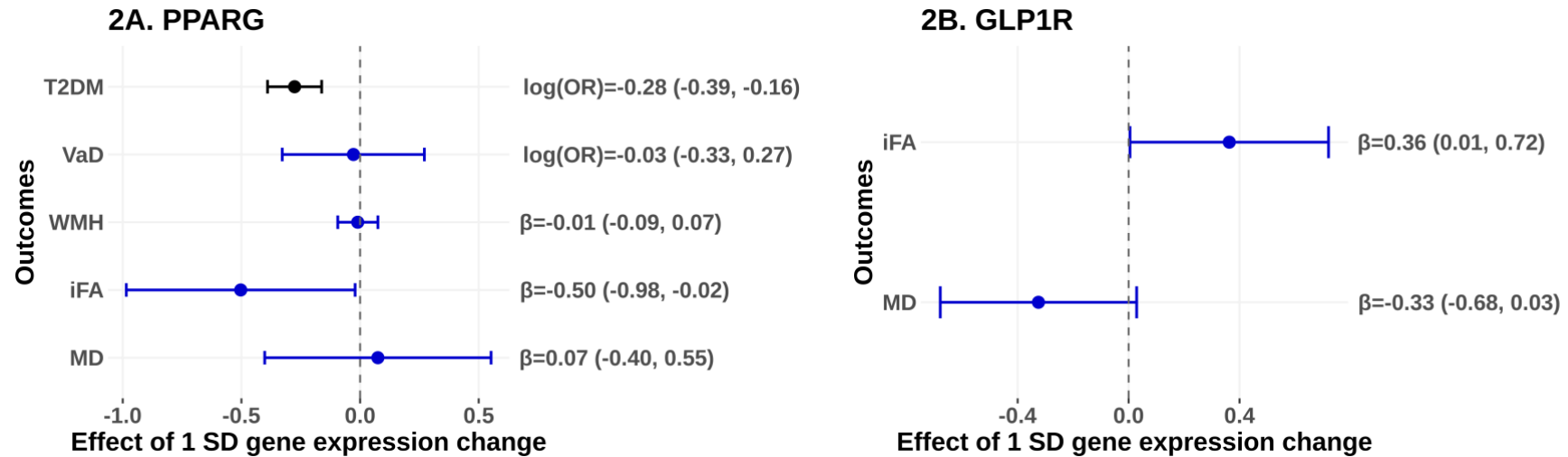

**Supplementary Figure 2. Secondary MR estimates of effects of modulating antidiabetic drug targets (*PPARG*, *GLP1R*) on T2DM risk and vascular dementia-related outcomes**

Results for the following targets are displayed as follows: (A) *PPARG* and (B) *GLP1R*. All estimates are expressed per 1-SD change in genetically predicted cortical gene expression and oriented to reflect the therapeutic mechanism of action of antidiabetic drugs acting on each target (agonism, antagonism, or inhibition). $\beta$ coefficients are plotted for continuous outcomes (WMH, iFA, MD), whereas log-odds are reported for binary outcomes (VaD and T2DM). The vertical dashed line represents the null.

Abbreviations: T2DM = type 2 diabetes mellitus; VaD = vascular dementia; WMH = white matter hyperintensities; iFA = inverse fractional anisotropy; MD = mean diffusivity; SD = standard deviation; MR = Mendelian randomization.

### STROBE-MR checklist

#### STROBE-MR checklist of recommended items to address in reports of Mendelian randomization studies

| Item No. | Section | Checklist item | Page No. | Relevant text from manuscript |
| --- | --- | --- | --- | --- |
| 1 | **TITLE and ABSTRACT** | Indicate Mendelian randomization (MR) as the study's design in the title and/or the abstract if that is a main purpose of the study | 1 + 2 | Title: "Evaluating Diabetes Drug Targets for Vascular Dementia Prevention Using Drug Target Mendelian Randomisation"<br>Abstract: "We applied two-sample, drug target Mendelian randomisation (MR) to test whether modulation of eight antidiabetic drug targets may alter VaD risk" |
| **INTRODUCTION** |  |  |  |  |
| 2 | **Background** | Explain the scientific background and rationale for the reported study. What is the exposure? Is a potential causal relationship between exposure and outcome plausible? Justify why MR is a helpful method to address the study question | 3 | "...several antidiabetic drug classes, including sodium-glucose cotransporter 2 (SGLT2) inhibitors, thiazolidinediones (TZDs), glucagon-like peptide-1 (GLP-1) receptor agonists, and dipeptidyl peptidase-4 (DPP-4) inhibitors, have been associated with lower dementia risk (Li et al., 2024; Tang et al., 2023). However, evidence remains inconsistent (Kuate Defo et al., 2024; Stefanou et al., 2026). Observational studies remain susceptible to residual confounding (Boyko, 2013; Eberly et al., 2021; Kornblith et al., 2022), while randomised controlled trial (RCT) evidence is limited and inconclusive (Li et al., 2024; Lin et al., 2023)."<br>"Drug target MR leverages genetic variants near genes encoding therapeutic targets to study the effects of pharmacological target modulation on disease outcomes (Gill et al., 2021). This approach can help prioritise candidates for drug repurposing, as therapeutic targets supported by human genetic evidence are approximately twice as likely to gain regulatory approval as those without such evidence (King et al., 2019)." |
| 3 | **Objectives** | State specific objectives clearly, including pre-specified causal hypotheses (if any). State that MR is a method that, under specific assumptions, intends to estimate causal effects | 3 | "We therefore used a two-sample drug target MR design to investigate whether genetically proxied modulation of eight diabetes drug targets influences risk of clinically diagnosed VaD and neuroimaging markers of cerebrovascular pathology." |
| **METHODS** |  |  |  |  |
| 4 | **Study design and data sources** | Present key elements of the study design early in the article. Consider including a table listing sources of data for all phases of the study. For each data source contributing to the analysis, describe the following: |  |  |
|  | a) | Setting: Describe the study design and the underlying population, if possible. Describe the setting, locations, and relevant dates, including periods of recruitment, exposure, follow-up, and data collection, when available. | 4 | "A two-sample, drug target MR framework (also known as *cis*-MR) was employed to evaluate whether modulation of diabetes drug targets influences the risk of VaD (Sanderson et al., 2022). For the primary analysis, *cis*-acting single nucleotide polymorphisms (SNPs) for each T2DM drug target were identified from a genome-wide association study (GWAS) of glycated haemoglobin (HbA1c) (Barton et al., 2021), a downstream biomarker indicative of target modulation across T2DM drugs." |
|  | b) | Participants: Give the eligibility criteria, and the sources and methods of selection of participants. Report the sample size, and whether any power or sample size calculations were carried out prior to the main analysis | Table 2 | We used existing GWAS summary statistics. Sample sizes and contributing cohorts are provided in Table 2. References to the original articles are also provided in Table 2 and within the manuscript, which provide full details on participant selection methods in detail. |
|  | c) | Describe measurement, quality control and selection of genetic variants | Table 2 | We used existing GWAS summary statistics. References to the original articles are also provided in Table 2 and within the manuscript, which describes measurement of genetic variants in detail. |
| | | | Pg. 7 | "For each diabetes drug target, a *cis*-region extending $\pm 500$ kb around the gene coding sequence was defined to minimise the risk of horizontal pleiotropy (Supplementary Table 1). SNPs within this region were retained if they met genome-wide significance ($p < 5 \times 10^{-8}$) with the exposure trait (HbA1c or eQTLs) ..." |
|  | d) | For each exposure, outcome, and other relevant variables, describe methods of assessment and diagnostic criteria for diseases | Table 2 | We used existing GWAS summary statistics. References to the original articles are also provided in Table 2 and within the manuscript, |

|  |  |  |  |
| --- | --- | --- | --- |
|  | e) Provide details of ethics committee approval and participant informed consent, if relevant | 5 | which describes measurement exposures and outcomes in detail.<br>“Ethical approval and participant consent for each contributing cohort were obtained in the original studies, as reported in the corresponding publications.” |
| 5 | **Assumptions**<br>Explicitly state the three core IV assumptions for the main analysis (relevance, independence and exclusion restriction) as well as assumptions for any additional or sensitivity analysis | 5, Supplementary Box 1, Figure 2 | “MR estimates are valid under the instrumental variable (IV) assumptions of relevance (IV1), independence from confounders (IV2), and absence of horizontal pleiotropy (IV3; Figure 2). Supplementary Box 1 outlines these assumptions and summarises the steps taken to minimise and evaluate potential violations.” |
| 6 | **Statistical methods: main analysis**<br>Describe statistical methods and statistics used |  |  |
|  | a) Describe how quantitative variables were handled in the analyses (i.e., scale, units, model) | 7 | “Results are reported as odds ratios (ORs) for binary outcomes and mean unit differences for continuous outcomes. Primary analyses were scaled per 0.1-SD genetically predicted reduction in transformed HbA1c attributable to target modulation, with estimates oriented to reflect the glucose-lowering action of diabetes drugs. Secondary analyses were scaled per 1-SD genetically predicted change in target gene expression, with effect directions aligned to the expected therapeutic action of each diabetes drug target (agonism, antagonism, or inhibition), ensuring genetically-proxied changes reflected the drugs’ mechanisms.” |
| | b) Describe how genetic variants were handled in the analyses and, if applicable, how their weights were selected | 7 | “For each diabetes drug target, a cis-region extending $\pm 500$ kb around the gene coding sequence ... SNPs within this region were retained if they met genome-wide significance ($p < 5 \times 10^{-8}$) with the exposure trait (HbA1c or eQTLs); if no genome-wide significant SNPs were available, a relaxed threshold of $p < 1 \times 10^{-5}$ was applied. To ensure independence among instruments, linkage disequilibrium (LD) clumping was performed using a 10,000 kb window and $r^2 < 0.001$ based on the 1000 Genomes EUR reference panel (The 1000 Genomes Project Consortium et al., 2015). The clumping p-value threshold ($p_1$) was matched to the SNP selection threshold applied for each |

|  |  |  |  |  |
| --- | --- | --- | --- | --- |
| | | | | analysis ($p < 5 \times 10^{-8}$ for genome-wide significance, or $p < 1 \times 10^{-5}$ where no genome-wide significant variants were available). For each drug target, a separate set of instruments was generated using this procedure from summary-level HbA1c and cortex-specific cis-eQTL data.” |
|  | c) Describe the MR estimator (e.g. two-stage least squares, Wald ratio) and related statistics. Detail the included covariates and, in case of two-sample MR, whether the same covariate set was used for adjustment in the two samples | 7 |  | “MR estimates were based on the Wald ratio (Sanderson et al., 2022) method for targets instrumented by a single SNP, and the inverse-variance weighted (IVW) method for targets with multiple instruments (Sanderson et al., 2022).” |
| | d) Explain how missing data were addressed | 6 | | “Where variants were unavailable in the T2DM dataset, proxy variants ($r^2 > 0.6$) were sought using the LDlink proxy tool (Machiela & Chanock, 2015), selecting the first available variant with the highest $r^2$ value. Full details of proxy selection methods and SNPs are available in the Supplementary Materials.” |
|  | e) If applicable, indicate how multiple testing was addressed | 7-8 |  | “No formal multiple-testing correction was applied. Bonferroni correction would be overly stringent in this setting, given multiple related drug targets across multiple related outcomes, raising potential false negatives. Instead, associations were interpreted cautiously, with careful consideration of confidence intervals and triangulation of supporting evidence.” |
| 7 | **Assessment of assumptions** | Describe any methods or prior knowledge used to assess the assumptions or justify their validity | 7, Supplementary Box 1 | “As all targets contained fewer than 3 instruments after data curation steps, sensitivity analyses that require 3 or more IVs were precluded. Instrument strength (IV1) was evaluated using F-statistics, with values over 10 taken to indicate sufficient relevance and low risk of weak-instrument bias (Burgess et al., 2011). Heterogeneity across variants (IV3) was evaluated using Cochran’s Q statistic when multiple instruments were available (Bowden et al., 2015).”<br>Supplementary Box 1 summarises the assumptions, methods used to mitigate their violation, and tests used to assess them or justify their validity |
| 8 | **Sensitivity analyses and additional analyses** | Describe any sensitivity analyses or additional analyses performed (e.g. comparison of effect estimates from different approaches, independent | 6 | “For the positive control outcome analysis, summary-level GWAS data for T2DM risk... Given |

replication, bias analytic techniques, validation of instruments, simulations)

that the selected agents are approved for the treatment of T2DM, genetic modulation of their corresponding targets should also be associated with T2DM risk, provided sufficient statistical power.”

7 “As all targets contained fewer than 3 instruments after data curation steps, sensitivity analyses that require 3 or more IVs were precluded. Instrument strength (IV1) was evaluated using F-statistics, with values over 10 taken to indicate sufficient relevance and low risk of weak-instrument bias (Burgess et al., 2011). Heterogeneity across variants (IV3) was evaluated using Cochran’s Q statistic when multiple instruments were available (Bowden et al., 2015).”

|  |  |  |  |
| --- | --- | --- | --- |
| 9 | **Software and pre-registration** |  |  |
|  | a) Name statistical software and package(s), including version and settings used | 7 | “All analyses were performed using R (version 4.4.3)” |
|  | b) State whether the study protocol and details were pre-registered (as well as when and where) | N/A |  |

### RESULTS

|  |  |  |  |
| --- | --- | --- | --- |
| 10 | **Descriptive data** |  |  |
|  | a) Report the numbers of individuals at each stage of included studies and reasons for exclusion. Consider use of a flow diagram | Table 2 | Table 2 summarises the number of participants contributing to each set of GWAS summary statistics used. |
|  | b) Report summary statistics for phenotypic exposure(s), outcome(s), and other relevant variables (e.g. means, SDs, proportions) | NA | The GWAS data used in this study is derived from the GWAS meta-analysis data of existing studies, so it is not reported. |
|  | c) If the data sources include meta-analyses of previous studies, provide the assessments of heterogeneity across these studies | NA | We did not perform a meta-analysis. We used existing GWAS summary statistics. |
|  | d) For two-sample MR: | 5 | i “All GWAS summary statistics were derived from individuals of European ancestry to minimise population stratification and confounding (i.e. violation of IV2).” |
|  | i. Provide justification of the similarity of the genetic variant-exposure associations between the exposure and outcome samples |  |  |
|  | ii. Provide information on the number of individuals who overlap between the exposure and outcome studies | Table 2 + 12 | ii Table 2 details the (partially overlapping) cohorts contributing to each set of GWAS summary statistics. “...partial sample overlap between exposure and outcome GWAS for some analyses, which may bias estimates and inflate Type I error |
|  |  |  | rates (Burgess et al., 2016). However, given adequate instrument F-statistics throughout, any bias is likely to be modest (Burgess et al., 2016; Sadreev et al., 2021)." |
| 11 | **Main results** |  |  |
|  | a) Report the associations between genetic variant and exposure, and between genetic variant and outcome, preferably on an interpretable scale | Supplementary Tables 3 & 4 | See the Supplementary Table 3 and Supplementary Table 4. |
|  | b) Report MR estimates of the relationship between exposure and outcome, and the measures of uncertainty from the MR analysis, on an interpretable scale, such as odds ratio or relative risk per SD difference | Pg. 9, Supplementary Tables 5 & 6, Figure 3, Supplementary Figures 1 + 2 | See the pages 10; "Primary Analysis: Targets Scaled to Effect on HbA1c.". As well as Supplementary Tables 5 and 6, and Figure 3 and 2, and Supplementary Figures 1 + 2. |
|  | c) If relevant, consider translating estimates of relative risk into absolute risk for a meaningful time period | NA | NA |
|  | d) Consider plots to visualize results (e.g. forest plot, scatterplot of associations between genetic variants and outcome versus between genetic variants and exposure) | Figure 3 and Supplementary Figures 1 + 2 | Please see Figure 3, and Supplementary Figures 1 + 2. |
| 12 | **Assessment of assumptions** |  |  |
|  | a) Report the assessment of the validity of the assumptions | Pg. 5, 9 & Supplementary Tables | Assessment of validity of the assumptions and attempt to mitigate violations:<br>"All instruments demonstrated sufficient strength (F-statistics > 10)" (IV1)<br>"All GWAS summary statistics were derived from individuals of European ancestry to minimise population stratification and confounding.." (IV2)<br>"...no evidence of heterogeneity (Cochran's Q, p > 0.050)" (IV3) |
| | b) Report any additional statistics (e.g., assessments of heterogeneity across genetic variants, such as $I^2$, Q statistic or E-value) | Pg. 9 & Supplementary Tables | "...no evidence of heterogeneity (Cochran's Q, p > 0.050), indicating consistent SNP-specific estimates." |
| 13 | **Sensitivity analyses and additional analyses** |  |  |
|  | a) Report any sensitivity analyses to assess the robustness of the main results to violations of the assumptions | 9 | "In the positive control outcome analyses, reduced T2DM risk was predicted for modulation of ..." |
|  |  | 7 | "As all targets contained fewer than 3 instruments after data curation steps, sensitivity analyses that require 3 or more IVs were precluded." |

|  |  |  |  |  |
| --- | --- | --- | --- | --- |
|  |  | b) Report results from other sensitivity analyses or additional analyses | Pg. 9 & Supplementary Materials and Tables | “Full results from the secondary analyses, using cortex-specific *cis*-eQTLs, are reported in the Supplementary Results and Supplementary Table 6...” |
|  |  | c) Report any assessment of direction of causal relationship (e.g., bidirectional MR) |  | NA |
|  |  | d) When relevant, report and compare with estimates from non-MR analyses |  | NA |
|  |  | e) Consider additional plots to visualize results (e.g., leave-one-out analyses) |  | NA |
| **DISCUSSION** |  |  |  |  |
| 14 | **Key results** | Summarize key results with reference to study objectives | 9-10 | “In this two-sample, drug target MR analysis, we found little evidence that modulation of currently licensed antidiabetic drug targets influences VaD risk or related neuroimaging outcomes - although a suggestive finding for DPP4 inhibition on VaD (combined with a lack of precision for that result) would warrant follow-up in future research” |
| 15 | **Limitations** | Discuss limitations of the study, taking into account the validity of the IV assumptions, other sources of potential bias, and imprecision. Discuss both direction and magnitude of any potential bias and any efforts to address them | 10-11 | “Several limitations warrant consideration...” |
| 16 | **Interpretation** |  |  |  |
|  |  | a) Meaning: Give a cautious overall interpretation of results in the context of their limitations and in comparison with other studies | 9-11 | “Collectively, these null findings provide genetically informed evidence that modulation of current diabetes drug targets is unlikely to yield clinically meaningful benefit for vascular dementia ...”, “...Previous evidence suggesting protective effects of specific T2DM drugs on VaD risk has largely been derived from individuals with diabetes, hyperglycaemia, or insulin resistance...”, and “...While this raises the possibility that PPARG activation could influence pathways relevant to white-matter integrity in VaD, interpretation remains tentative...”. |
|  |  | b) Mechanism: Discuss underlying biological mechanisms that could drive a potential causal relationship between the investigated exposure and the outcome, and whether the gene-environment equivalence assumption is reasonable. Use causal language carefully, clarifying that IV estimates may provide causal effects only under certain assumptions | 10 | We include limited discussion of mechanisms, given the generally null findings.<br>“Overall, our findings provide little genetic evidence supporting currently licensed antidiabetic drug targets as priorities for vascular dementia prevention. These results are informative for |
|  |  |  |  | therapeutic prioritisation because they suggest that several widely discussed diabetes drug targets are unlikely to exert large protective effects on VaD risk, while identifying *DPP4* inhibition as a target warranting further investigation” |
|  |  | c) Clinical relevance: Discuss whether the results have clinical or public policy relevance, and to what extent they inform effect sizes of possible interventions | 10-12 | “...genetically informed null findings are valuable for therapeutic prioritisation because they can identify targets that are unlikely to yield clinically meaningful benefits, thereby reducing investment in costly and lengthy dementia prevention trials with a low probability of success...” |
| 17 | **Generalizability** | Discuss the generalizability of the study results (a) to other populations, (b) across other exposure periods/timings, and (c) across other levels of exposure | 11 | “...our results reflect target-specific effects of perturbing specific molecular targets, rather than the effects of administering the corresponding drug ...”<br>“...genetic variants proxy lifelong perturbation of drug targets and therefore most closely resemble the effects of long-term target modulation. These estimates may not correspond to the effects of pharmacological treatment initiated later in life or administered over shorter durations...”<br>“...restricting analyses to European ancestry likely reduced bias from population stratification (minimising potential violations of IV2; Supplementary Box 1). However, the generalisability of findings to other ancestries remains uncertain...” |
| **OTHER INFORMATION** |  |  |  |  |
| 18 | **Funding** | Describe sources of funding and the role of funders in the present study and, if applicable, sources of funding for the databases and original study or studies on which the present study is based | 12 | “ELA is supported by a UKRI Future Leaders Fellowship (MR/W011581/1). DMW is supported by an Alzheimer’s Research UK Senior Fellowship (ARUK-SRF2023B-008). The funders had no role in study design, data collection and analysis, decision to publish or preparation of the manuscript.” |
| 19 | **Data and data sharing** | Provide the data used to perform all analyses or report where and how the data can be accessed, and reference these sources in the article. Provide the statistical code needed to reproduce the results in the article, or report whether the code is publicly accessible and if so, where | 12 | “All summary statistics were obtained from publicly available datasets or directly from the corresponding authors...” |
| 20 | **Conflicts of Interest** | All authors should declare all potential conflicts of interest | 12 | “The authors declare no conflicts of interest” |

This checklist is copyrighted by the Equator Network under the Creative Commons Attribution 3.0 Unported (CC BY 3.0) license.

*International Journal of Epidemiology*, 46(6), 1734–1739.

https://doi.org/10.1093/ije/dyx034
